# Beyond Qualification: a Video-Stimulated Interview Study on What Group Discussion of Audit and Feedback Adds to Professional Development in General Practice

**DOI:** 10.1101/2025.06.20.25329918

**Authors:** Dorien van der Winden, Mana Nasori, Marianne Mak-van der Vossen, Nynke van Dijk, Marije van Braak, Jettie Bont, Mechteld Visser

**Author notes:** **Corresponding author:** Dorien van der Winden, Amsterdam UMC department of General Pratice, Meibergdreef 15, 1105AZ, Amsterdam, The Netherlands,. This project was funded by the department of General Practice at the Amsterdam UMC. No external funding was received.

## Abstract

**Introduction:** Modern healthcare demands continuing professional development (CPD) that bridges learning and practice and motivates health professionals. Current CPD activities often prioritize qualification, while socialization and subjectification purposes are considered vital in other educational contexts and could help to foster dynamic learning. Group discussion of audit and feedback (A&F) is a form of CPD that is used increasingly, and that may fulfill these needs. However, little is known about how this approach creates learning opportunities or what the group dynamic contributes.

**Method:** In this video-stimulated interview study we explored how a group of experienced general practitioners perceive the value of group discussion of A&F in their professional development, employing a constructivist paradigm. We first inductively analyzed our findings, using Thematic Analysis. A second deductive analysis followed, using Biesta’s concept of the three purposes of education (qualification, socialization and subjectification) as framework.

**Results:** According to our participants, group discussion of A&F allows for a reflective process that is formed by group discussion. The group helps deepen reflection and assists with the forming of individual and collective opinions. The meetings enhanced motivation for both individual and collective learning. Key conditions included a safe learning environment and a high level of enjoyability. The group meetings offered opportunities for all three purposes of education.

**Discussion:** Group discussion adds value to individual A&F by offering room for socialization and subjectification, as well as classic qualification purposes. It thereby offers a future proof form of CPD that could improve quality of healthcare and stimulate lifelong learning.

## Introduction

The quickly changing landscape of healthcare asks for health professionals that are highly adaptive and continuously work on developing themselves and their practice. Strong strategies for continuing professional development (CPD) are essential to stimulate lifelong learning.(1, 2) Yet, existing CPD methods are being critiqued for often being too focused on ‘getting credits’ and for not being properly anchored in practice.(3) This results in lack of engagement with CPD and potentially impairs lifelong learning.(2) We therefore need CPD methods that link learning to practice and encourage engagement. Group discussion of audit and feedback (A&F) combines CPD with quality improvement (QI), is strongly rooted in practice and could possibly foresee in this need for a different kind of CPD activity.(4)

A&F interventions are traditionally used in QI settings.(5) In A&F interventions, health data (audit), is fed back to medical professionals in order to improve care.(6) The most recent Cochrane review states that A&F activities have mild to moderate effects.(7, 8) Adding a social factor, such as group discussion, seems to enhance the effect.(9-11) Currently, A&F is implemented in a CPD context more often, meaning it focuses on reflection and learning. An example is group discussion of A&F, where health data reports are discussed with a group of peers.(4, 12) Since most research on A&F interventions has been done with a QI lens, focusing on outcomes and effect measurement, it is largely unknown how group discussion of A&F works in a CPD setting. To gain more insight in how group discussion of A&F leads to reflection and offers learning opportunities, adopting an educational perspective would be helpful.

Biesta’s work on the purposes of education offers an interesting education philosophical take on reflection and learning processes. Biesta states that education should not only focus on qualification (transition of knowledge and skills), but also on socialization (understanding and internalizing the values of the profession) and subjectification (relating oneself to the profession and finding your own position).(13, 14) Whereas Biesta’s insights were formerly used mostly in primary and secondary education, they are currently also applied in vocational and (post)graduate training.(15) For example, the new curriculum for general practice in the Netherlands focuses highly on integrating ‘getting qualified’ with socialization and subjectification.(16) In CPD settings though, the focus of both researchers and professionals is still very much on maintaining qualification.(2) This focus stems partly from the design of reregistration systems worldwide, that are based mainly on keeping oneself qualified for the job.(2, 17, 18) There is little attention for socialization and subjectification, while it seems highly likely that these goal domains also remain relevant after completion of postgraduate training.(17, 19)

In this study we adopted an educational perspective and studied a best practice initiative of group discussion of A&F to explore the following research questions:

1. How does group discussion of A&F offer learning opportunities, according to participants?
2. How do participants believe these meetings add value to their professional development?
3. What factors contribute to establishing a beneficial meeting?

We then explore how these findings relate to Biesta’s purposes of education: qualification, socialization and subjectification, with the purpose of informing future initiatives in practice as well as future research.

## Methods

### Study design and theory

Within a constructivist paradigm, we chose a video stimulated reflective interview format.(20) This suited our objective to explore ideas of participants while sticking closely to what actually happened in the meetings. Adopting an inductive set-up, we decided not to define learning in advance, but to ask our participants what they found valuable about group discussion of A&F and what they gained from it. This fits Biesta’s ideas on learning: the cognitive process of learning itself is impossible to explicate, while more insight is offered by exploring the presence of valuable moments within an educational activity and how this adds to the development of participants.(13)

### Reflexivity

A constructivist paradigm sees knowledge as actively constructed and cocreated as a result of human interactions and relationships. To inform our readers of our own stance, see Table 1 for the authors’ backgrounds. Additionally, all have experience in performing and receiving (medical) education and researching it. The researchers are affiliated with a large general practice research and training institute. The overarching research project came to exist through a request posited by a group of local general practitioners (GPs), asking for a scientific framework for their CPD activities. This stimulated us to keep our research aim closely connected to practice, while also furthering theory. DW’s own experience in general practice offered the opportunity to stick closely to the context. However, it may have also created a narrower mindset, which is why adding researchers with different backgrounds was important. We maintained a reflexive stance throughout the research process by frequently discussing our positionality and critically questioning each other on its implications.

**Table 1.** Background of the authors.

| Author | Background |
| --- | --- |
| DW | GP in final stages of training<br>PhD student in medical education |
| MN | Health policy<br>PhD student in patient participation |
| MM | GP<br>Medical educator / Assistant Professor in HPE |
| ND | Professor in medical education<br>Dean of HPE faculty |
| MB | Assistant Professor in communication studies<br>Language specialist |
| JB | GP<br>Professor in general practice |
| MV | Senior researcher in Medical Education<br>Cognitive psychologist by training<br>Quality expert |

### Context

We conducted our study within a general practice setting in the Netherlands. Dutch GPs need to reregister every five years.(21) For reregistration, GPs must dedicate at least 10 hours per five years to peer-review CPD activities, such as group discussion of A&F. To facilitate peer-review activities, GPs participate in voluntarily formed groups. Group dynamics vary significantly among different groups, as some groups are collaborating intensively in several respects, while other groups only come together for the obligatory hours of peer-review.

### Participants and set-up of meetings

We purposively recruited an established group of GPs who had been participating satisfyingly in group discussion of A&F for several years. Studying this “best practice” group fitted our research questions: it allowed us to broaden our understanding of how group discussion can be valuable and offers insight in what factors are pivotal for this. This group, consisting of 12 GPs, developed their own format for their meetings. The subject of the meeting is determined by the members of the group. A short questionnaire on the subject is sent out beforehand, covering knowledge items and self-report on how well a subject is mastered. The meeting contains a presentation by a group member on the subject, followed by a discussion of A&F reports on practice level. In some meetings they are joined by an external expert on the discussed subject (a specialised GP or medical specialist). The meeting ends with stating points of improvement by each member, which they revisit after a year, to map progress.

### Data collection and interviewing

Data collection and analysis was performed iteratively. We observed and video recorded two regular A&F group meetings. After the first meeting the video material was viewed by two individual researchers (DW and MV) with the purpose of identifying key learning opportunities in the meeting. Videos were then watched together by DW and MV, reaching consensus on which opportunities to discuss within the interviews that would follow. When necessary, a third researcher (ND) was consulted. After this, we conducted a round of individual video-stimulated interviews. Participants were selected based on their availability, preferably within a three-week window of the meeting.

Semi-structured interviews were performed by DW and MV in a location chosen by the participant. Participants were asked if they recalled moments they wanted to review. Because all participants indicated that they would not have time to review the footage themselves, we used video fragments that were selected by the researchers. For each participant, we used a basic topic list that was individually constructed, based on the video material, structuring the interview while also offering space for questions that arose.[See additional materials 1](22) The interviews were audio-recorded and transcribed verbatim. For the second meeting the same format applied.

### Analysis

We used Thematic analysis to analyse transcripts of the interviews.[ref Braun Clarke 2022] Its flexible and overarching approach suited our broad research objectives. We used MAXQDA software.(23) Analysis was done by two researchers independently (DW and MN), consulting each other after each interview and iteratively building on a shared coding tree. In this first inductive analysis, open coding was used. MV was consulted after every two interviews. Consensus was reached on a final coding tree, through discussion.[see additional materials 2] After analysing the second round of interviews, we evaluated findings with the whole research team and concluded that our data was sufficiently rich to answer our research questions.

After inductively analysing all transcripts, the research team explored potential theories to contextualize the findings. Biesta’s work on the purposes of education seemed to fit how our participants described their learning experiences.(13, 14) DW and MM went back to the raw data and deductively analysed the original data, using Biesta’s three purposes as a priori codes, forming our framework. After discussing the results with two additional researchers (MN and MV), the final deductive coding tree was constructed.[see additional materials 3] Consensus about the final results was reached within the full research team.

### Ethical consideration

The Medical Ethical Board of Amsterdam UMC regarded the study protocol not to be subject to the Medical Research Involving Human Subjects Act. Consequently, they provided a waiver (numbered A1 19.333). All data obtained within this study was processed and stored according to the General Data Protection Regulation and according to the Amsterdam UMC Clinical Research Unit procedures. In addition to an information meeting on the study’s purpose and privacy concerns, all eligible participants were sent an information letter. All participants individually gave informed consent for observation and video recording of the group meetings. Participants who agreed to observations and video recordings of the meetings were allowed to opt out of participating in the interviews.

## Results

Video recordings were made at two A&F group discussion meetings. In the first meeting a member of the group provided the knowledge background, while in the second meeting a content-related expert was present. Eight interviews were held with seven participants. The moderator of the group was interviewed twice, i.e. after each meeting. See Table 2 for participant characteristics.

**Table 2.**
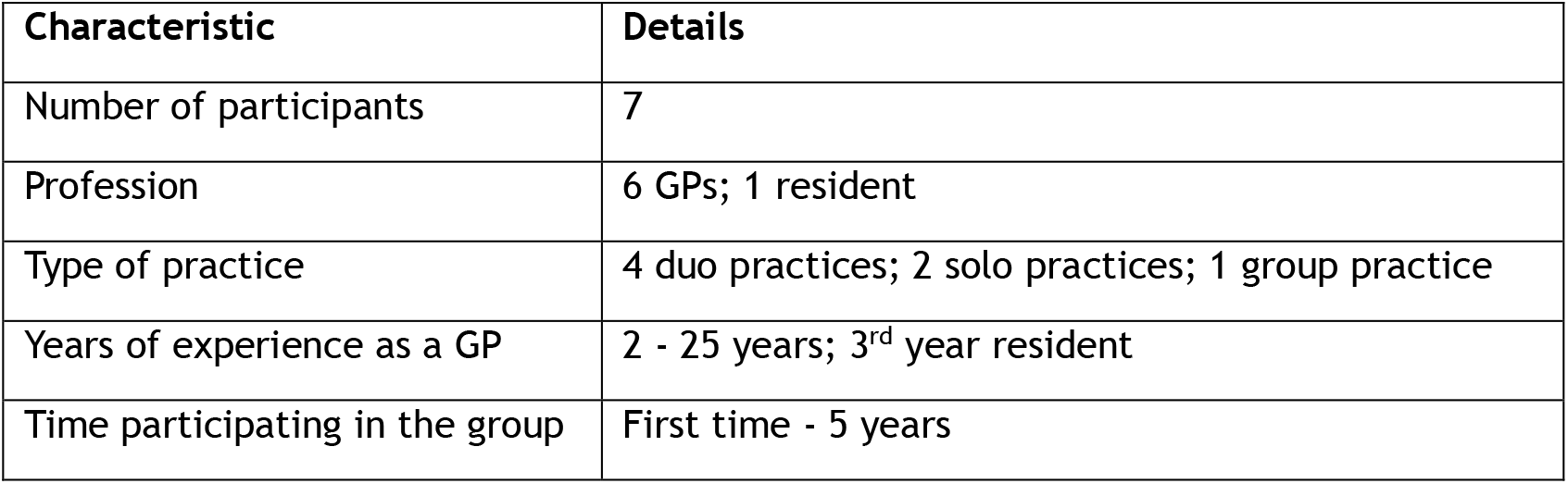
Participant characteristics.

In the following paragraphs we first describe our inductive findings, answering our research questions. Subsequently, we depict the deductive results, indicating how qualification, socialization and subjectification are present in our findings.

### Group discussion deepens reflection

Participants explained that the group aspect of the meetings was particularly valuable. The A&F reports were said to connect the meeting to daily practice and were used to initiate reflection. Participants explained that, when confronted with numbers that stand out from the rest in a negative sense, others helped them to reflect by asking questions and thinking along, prompting questions such as: why are my numbers so different from the others? Do I organize my practice differently? Do I have different values or a different idea on what is proper care? This reflection led to important insights, aiding in identifying potential issues, areas for improvement, or aspects that required further investigation. The group was considered assistive to establish *a more meaningful level of reflection*, harder to reach individually.(Table 3, quote 1)

**Table 3.** Quotes 1 and 2: Deepening of reflection.

|  |  |  |
| --- | --- | --- |
| 1. | *‘On the other hand, there are topics where you can have strong opinions, and as a group, you might be able to question each other, so that you get to a deeper layer of “Oh, what are actually my assumptions about this, and what effect does that have?”’* | Participant 1 |
| 2. | *“Yes, on your own, you look at things in a certain way. I do my medicine the way I always do it, so I also think I look at those numbers the way I would usually look at them. But when others say something about it, you do hear things from outside your normal way of working.”* | Participant 3 |

Also, participants described that even if they were not the one actively reflecting, it was still helpful to hear others talk about the subject and reflect on their practice. This offered perspective on participants own situation or functioning. It also contributed to forming new ideas and opinions of their own. Oppositely, when doing relatively well compared to the others, this enhanced participants confidence and even resulted in more job satisfaction.(table 3, quote 2)

### Group discussion induces both individual and collective learning

Also, the group process was described to be pivotal for *forming both individual and collective opinions*, which happened by discussing the subject and hearing ideas and opinions of others.

Subjects that especially benefitted from group discussion, were those for which there is limited evidence, or on which professional guidelines leave room for interpretation: participants felt the need to discuss these subjects with their colleagues and *form a collective opinion* on how to tackle these subjects. Since in daily practice they often work solo, these group meetings filled a gap, in this respect. The phenomenon of collective learning brought joy and resulted in motivation to perform their profession and to keep learning.(table 4, quote 3)

**Table 4.** Quote 3: Inducing individual and collective learning.

|  |  |  |
| --- | --- | --- |
| 3. | *“So, by grappling with the material, by discussing or questioning each other, I understand it... Yes, or ‘understand it,’ I do, but it also makes me remember it better. It also offers me pleasure to go do something with it again.”* | Participant 7 |

### Group discussion prepares for collective action

Another effect of the group meetings that was described, is that participants identify common problems in the meetings, often caused by working in the same setting. These could be solved by *collective action*. A mentioned example was problems with referral of patients to the local hospital. Participants said the meeting empowered them to take collective action, while they would not have felt able to do so individually.

### Key figures create a safe learning environment

The safety of the learning environment was considered essential because it created space for vulnerability, which our participants deemed necessary for reflection and learning. Participants described this as a ‘there is no right or wrong’ atmosphere. This *safe learning environment* was actively created by key figures, such as the moderator or other experienced GPs in the group, making themselves vulnerable and openly showing their insecurities. This encouraged other group members also feeling safe to share their own experiences. At the same time, the presence of these experienced role models could be intimidating for some participants: putting them on a pedestal could also impair the willingness of participants to share their insecurities.(table 6, quote 5 and 6)

**Table 5.** Quote 4: Collective action.

|  |  |  |
| --- | --- | --- |
| 4. | *“...but then a second goal of such a community health group is democratization. If something is felt in the field, it must be communicated to hierarchies above this group, so that it is actually addressed there. ... So now you need to try to generate enough volume for the issue and then push it forcefully to the right place. Yes, that then becomes a bit of a political piece. That can also be the outcome of the [group discussion of AF] session. Yes.”* | Participant 1 |

**Table 6.** Quote 5 and 6.

|  |  |  |
| --- | --- | --- |
| 5. | *'At some point, there were names of practices involved . Yes, then you have no choice but to expose yourself. But even then, there's an atmosphere of "there's no right or wrong".'* | Participant 5 |
| 6. | *'You asked: 'What's it like, being in such a group?' The fact that I don't mind Ma. asking me that... It's also a very safe group. That absolutely helps.'* | Participant 2 |

### Enjoyment is a prerogative

Enjoyment was considered crucial for the meetings to continue. Participants described enjoying the meetings, which was a key reason to join them and to keep showing up. The moderator considered making the meetings enjoyable a large part of her assignment. This also influenced her behavior as moderator: ensuring enough laughs and adding jokes when things got heated or someone was called out. This was connected to the non-compulsory nature of the meetings but was also considered helpful for learning.(table 7, quote 7)

**Table 7.** Quote 7: Enjoyment.

|  |  |  |
| --- | --- | --- |
| 7. | *'Everyone should enjoy it a bit and has to want to come , because I have no other leverage, so I just have to make it fun.'* | Participant 1 |

### Qualification, socialization and subjectification

All three functions of education could be recognized in our participants descriptions of learning from A&F group meetings. *Qualification* was present as looking at the numbers and seeing how well you were performing (as a means of assessment) and in the information/ knowledge sharing part of the meetings (where either the expert or one of the members of the group explained the guideline/status quo on the subject of each meeting).(table 8, quote 8)

**Table 8.** Qualification, Socialization and Subjectification.

|  |  |  |
| --- | --- | --- |
| 8. | *'Most of them are really experienced doctors, and they've been working in a certain way for twenty years, but they also have an exceptionally good clinical eye and a lot of experience. Look, working the same way for twenty years isn't good, of course. You must keep up with the times, and you need to be flexible. That's definitely what's being discussed properly here.'* | Participant 3 |
| 9. | *'It's really about searching together for "how do we do this now?" ... It's definitely about searching together for "what motivates us to do it the way we're doing it?"'* | *Participant 2* |
| 10. | *'Of course, there are topics where there is much more of a norm. Sometimes, it's like everyone is deviating from the norm, like with oxycodone, and you can still learn from each other in how you deal with it and the solutions you're going to look for, talking about it together, like "who would know more about this, who could we possibly refer to, how do we prevent this exactly, what's our first choice pain relief," giving that some structure. Then you're searching together, and you have consensus on the norm...'* | *Participant 1* |
| 11. | *'What do you do in that consultation room? You must think about that. And then you need to think further; the next step is "how do you want to do it?"'* | *Participant 2* |

**Table 9.** Conflicting purposes.

|  |  |  |
| --- | --- | --- |
| 12. | *'...and that's also generally the flaw of bringing in an expert, because it's much easier to just pick up those pieces of knowledge again than to reflect.'* | *Participant 1* |

*Socialization* was recognized in participants description of what group discussion led to: when discussing subjects for which there is little evidence, or where guidelines leave room for interpretation, this resulted in a shared construction of values, norms and ideas on what they believe good GP care is. This was considered one of the main functions of the meetings: by discussing subjects, a collective concept formed around the discussed subject.(table 8, quote 9 and 10)

*Subjectification* was clearly identified as both an objective for attending the meetings and as a result of the meeting. Through the process of discussion and forming opinions as a group, participants discovered their own position in relation to the discussed subject. This was often prolonged after the meeting: back in their practices this thinking process continued, for example through conversations with their direct colleagues. Additionally, this resulted in relating themselves to their profession: what kind of doctor do I want to be? What do I find important? What do I believe is the right care, and how does this become visible in my daily doctoring?(table 8, quote 11)

### Purposes could be conflicting

The meetings covered all three purposes, although among the participants some conflicting wishes were present. Participants that favored qualification as the main purpose, expressed a strong wish for an expert to be present. Opposingly, a participant that recognized the priority of subjectification, said that the presence of an expert can impair discussion among the group, overshadowing the group process. Since the expert is there to explain what is right and what is wrong, this allowed the group less room for discussion and to explore their own ideas.

**Figure 1.**
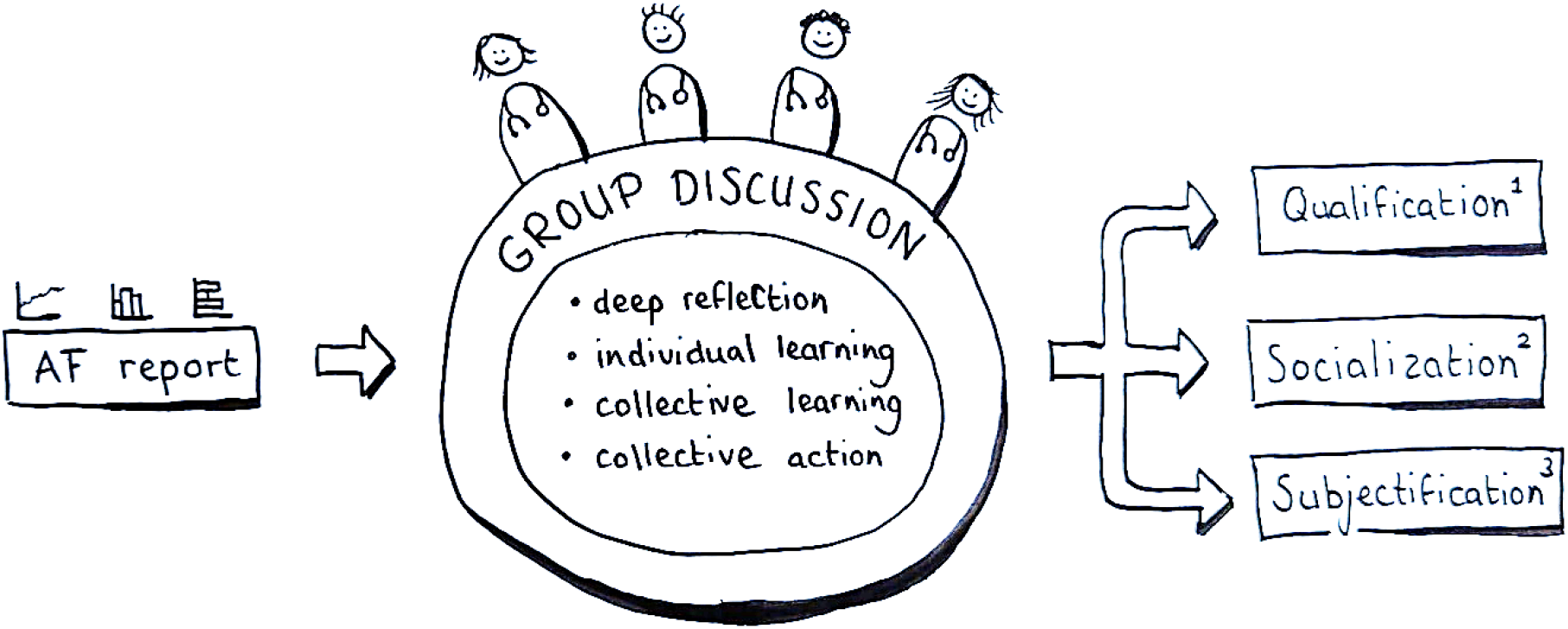
Visual summary of results; 1. Qualification, understood as: transition of knowledge and skills; 2. Socialization, understood as: understanding and internalizing the values of the profession; 3. Subjectification, understood as: relating oneself to the profession and finding your own position.

## Discussion

In this article we set out to understand how participants believe group discussion of A&F offers learning opportunities and how these meetings add value to professional development of GPs. Our results show that the A&F reports serve as the start of a reflective process that is shaped by group discussion. The group helps to *deepen reflection* and with forming *individual and collective opinions*, especially on subjects where less evidence is available. The meetings stimulated motivation for individual and collective learning. Key conditions to make these meetings valuable were the presence of a *safe learning environment* and a high level of *enjoyability*. The group meetings offered opportunities for all three domains of purpose of education: *qualification, socialization* and *subjectification*.

### The added value of the group

As we set out to broaden our understanding of what the group adds, when studying A&F, we found that the added value is most present within the process of discussing the A&F together: learning opportunities occur within and through discussion. This resonates with the broader educational literature on social learning, starting with Bandura’s theories, published in the 1970’s.(24) This theory describes learning as an inherently social process: learning begins with looking at peers. These principles are widely used in primary, secondary and undergraduate education. Our findings show that it also stands firm in this CPD setting, even though traditionally CPD is often an individualistic undertaking. Our results show that the aspect of learning together with colleagues is especially appreciated and even motivates participants to keep learning, both individually and collectively. This insight within the CPD context is an important one: the focus of CPD should, at least in part, shift from the individual to the collective and make more use of group interventions such as group discussion of A&F.

### Learning together stimulates collaboration

Our results show that these group meetings create the opportunity for collective action. This means using more group based CPD activities could also stimulate collaboration and understanding between professionals. Both are pivotal for modern high-level healthcare.(25, 26) For general practice this stimulation of collaboration is especially relevant, as daily practice of GPs is highly based around the individual doctor. In the meetings our participants found space to come together, learn together and to gain a better understanding of their own work and that of others.

### Beyond qualification

Coming together, forming collective insights and understanding your individual stance on this, was one of the much-appreciated gains of group discussion of A&F, as indicated by our participants. These processes can be understood as forms of socialization and subjectification. All the while, participants feel that qualification should still be strongly represented. This resonates with Biesta’s findings that all three purposes need attention.(13) Even though the purposes are not traditionally used for a CPD setting, our study indicates that in group discussion of A&F this combination seems to be a strength. This fits recent findings that a broader conceptualization of lifelong learning and CPD is necessary, focusing not only on qualification, but additionally offering space for socialization and subjectification.(2, 19) In our study we showcased this by providing a real-life best practice example of how combining the three purposes helps professionals in their development. We thereby add to the evidence that seeing CPD in a broader context and acknowledging the three functions equally, could possibly help professionals to navigate the changing landscape of healthcare.(2, 17, 19)

### Socialization and subjectification examined

Looking closer at socialization, we realize that it is important to clearly define the concept of socialization. Our results show that, especially lesser researched subjects and subjects where guidelines leave room for interpretation, invoke discussion. This discussion resulted in forming an opinion as a group and deciding upon norms and values (‘what do we believe good/proper care is?’). We argue that this is what socialization could look like in a CPD setting: not only conforming to existing norms and values of the profession, but also forming new ones. Socialization is often defined as ‘making the norms and values of the group your own’. We would like to plead for a broader definition, when used in a CPD context: socialization is not only about making the norms and values of the group your own, but also about forming new norms and values together, as a group. This broader definition fits Brown and Finns concept of ‘post-functionalist’ socialization, and reads as a more fluid definition of socialization: one where the norms and values are only partly set, and can still be shaped.(27) We see in our results that through group discussion, socialization and subjectification seem to go ‘hand in hand’: through discussion a common denominator is formed, while participants simultaneously start positioning themselves to this common denominator as an individual.

### Safe learning environment and enjoyment

Applying Biesta’s insights poses the question of how to interpret the importance of a safe learning climate, and of enjoyment, in this light. The need for a safe learning environment is widely spread throughout pedagogical and educational literature, also fitting Biesta’s description of the need for ‘objective spaces’ to find answers.(13) This objective space, where learners feel safe to explore the world, to learn by discovering and making mistakes, is vital to learning, or rather, to facilitate qualification, socialization and subjectification. In Biesta’s theory, the teacher is an important player in providing this space. In group discussion of A&F, we believe that the moderator in collaboration with the group can provide this ‘objective space’ by ensuring vulnerability.

Enjoyment contributed to a more engaging and productive learning environment. However, focusing on enjoyment poses a danger: if all friction is avoided, learning could be impaired. According to Biesta this is exactly one of the dangers of modern education: continuously trying to make learning fun, results in missing important learning opportunities.(14) Learning cannot always be pleasant: sometimes learning is hard, or even unpleasant. Our data show that the moderator plays a pivotal role in both the safety of the learning environment and properly balancing enjoyment. We therefore recommend to carefully select the moderator, who is preferably a key figure from within the group, leading firmly but also showing and allowing vulnerability in a role model position.

### Strengths and limitations

To our knowledge, our study is the first to focus from an educational perspective on the process of group discussion of A&F. Our use of educational theory is a strength and offers possibilities for further research in wider contexts. This study adds to the knowledge on group discussion of A&F and is the first to gain an understanding of the participants’ perspectives by using a video-stimulated interviewing setup. This makes the insights gained from the interviews strongly connected to what actually happens during the meetings. Even so, our study is one of modest proportions: it is exploratory and participants are all part of only one CPD group. Our findings require further research to see whether they resonate with various groups in various contexts. Additionally, studying a best practice group means that we interviewed people who are experienced with, and mostly enthusiastic about, the format. This fitted our purpose of broadening our understanding of what we could gain by adding group discussion to A&F interventions, but it is good to keep in mind that it does not shine light on potential problems and risks.

### Implication for design, practice and future research

Understanding that group discussion of A&F is not just about qualification, but also offers space for socialization and subjectification, can help in designing, evaluating and researching this type of intervention more effectively. All three purposes in mind, we understand better when and how to use group discussion of A&F. When researching this type of group intervention of A&F, we need to rethink our setups: we will need to map outcomes and effects beyond just qualification, and closely examine the process, as we get a better understanding of what health professionals can gain from it.

## Conclusion

Concluding, group discussion of A&F adds value to professional development in general practice by deepening reflection and allowing for individual as well as collective learning and action. Group discussion adds value to individual A&F interventions by offering room for socialization and subjectification, as well as classic qualification purposes. The group process motivates and stimulates collaboration. It thereby offers a future proof form of CPD that could improve quality of healthcare and stimulate lifelong learning.

## Data Availability

All data produced in the present study are available upon reasonable request to the authors.

## Acknowledgements

We thank all our participants for allowing us to observe their shared learning environment and for their reflectiveness on their learning processes.

## Data accessibility

Due to privacy concerns our original data are available through the corresponding author upon request. In our additional materials you can find:

1. Additional materials 1 Topic List – English
2. Additional materials 2 Codebook Inductive Analysis
3. Additional materials 3 Codebook Deductive Analysis

