## Additional materials 1 for "Beyond Qualification: a Video-Stimulated Interview Study on What Group Discussion of Audit and Feedback Adds to Professional Development in General Practice"

**Topic List – Video Stimulated Reflection Interview Study – Audit and Feedback Group Discussion**

**Introduction:** We are interested in what exactly happens when audit and feedback information is discussed in a group. Do you learn more from this than when you look at it alone? What makes the difference? As researchers, we could observe such a group meeting and form an idea about it, but in this study, we are curious about how the participants themselves view this. We want to open this "black box" a little further with your help.

**To begin, a few general questions:**

- How long have you been a general practitioner (GP)?
- How long have you been working in your current practice?
- Since when have you been part of this review group?

On [date], the neighborhood group meeting was held on [topic]. As you know, we filmed the session. Using a few video clips, we would like to reflect with you on what exactly happens during such a group meeting. We are looking for your thoughts on this and would like to discover together how this process works.

First, I would like to ask if there are any specific moments you would like to revisit? I will try to find those moments in the video material. Which moments are those? What makes you want to revisit these moments?

**What do you think of review group meetings like this, where you all look at the data together?** Does anything different happen compared to when you look at your data alone? What makes it different? How does the group contribute (or not) to this process? Are there any moments from the last meeting where you think the group really added something? Can you recall such a moment?

We have a few video clips we would like to show you.

**Questions for the video clips:**

- You said here … (neutral observation)
- What is happening here?
- You seem to … (observation)
- What made you do this? What did you achieve with this?
- What do you think happened here? What was your opinion about this?
- What is the influence of the group on what happened here?
- What is the outcome of what happened here?

***Note:*** *Keep in mind: where does the group really have an impact? Which phase of learning is the interviewee referring to? Is it about acquiring knowledge / internalizing / gaining insight / motivation / intention to change / self-efficacy / generating ideas on how to implement it?*
